# Lipoprotein(a) is Associated with Increased Low-Density Plaque Volume

**DOI:** 10.1101/2024.07.18.24310539

**Authors:** Rebecca Fisher, Nick Nurmohamed, Edward A. Fisher, Melissa Aquino, James P. Earls, James K. Min, Chen Gurevitz, Waqas A. Malick, M. Robert Peters, Sascha N. Goonewardena, Robert S. Rosenson

## Abstract

**BACKGROUND:** Lipoprotein(a) [Lp(a)] is an inherited risk factor for cardiovascular disease that is accompanied by a more severe coronary artery disease (CAD) phenotype and a higher risk for events. The objective of this study is to clarify the association between Lp(a) and coronary plaque characteristics in asymptomatic patients.

**METHODS:** 373 consecutive asymptomatic patients were evaluated for primary prevention of CAD. Artificial intelligence quantitative coronary CTA (AI-QCT) was used to investigate the relationship between Lp(a) and coronary plaque characteristics. Multivariable linear regression adjusted for CAD risk factors (age, sex, race, diabetes, smoking), statin use, and body mass index were used to analyze associations between the Lp(a) (by quintile), high sensitivity C-reactive protein (hsCRP), coronary artery calcium (CAC) score, and AI-QCT findings. AI-QCT findings were defined as low-density non-calcified plaque volume (LD-NCPV).

**RESULTS:** The mean age was 56.2±8.9 years, 71.6% were male, and 54.2% were taking statin therapy. Median LDL-C was 103(72,136)mg/dL, median Lp(a) was 31(11, 89)nmol/L, median Lp(a) corrected LDL-C was 101(64, 131)mg/dL. Median hsCRP levels were 0.8(0.4, 1.8)mg/L. Median CAC levels were 6.0(0.0,110.0). There was no association between Lp(a) concentrations and CAC(P=0.281). After adjustment for CAD risk factors, every quintile of Lp(a) increase was associated with a 0.4% increase in LD-NCPV(P=0.039). The inclusion of hsCRP to the models had no significant effect on LD-NCPV.

**CONCLUSIONS:** Higher Lp(a) concentrations in asymptomatic patients are significantly associated with increased low-density non-calcified plaque volume.

**Clinical Perspective:** Lp(a) is a risk marker for early-onset coronary heart disease events. Early detection of vulnerable patients is critical to mitigating this risk that may be inadequately captured by the coronary artery calcium score. Low-density non-calcified plaque quantification by coronary computerized tomography is an approach that may be more suitable to assess risk in patients with high Lp(a) levels.

## INTRODUCTION

Elevated lipoprotein (a) [Lp(a)] is an inherited trait that is associated with an increased risk of atherosclerotic cardiovascular disease (ASCVD) and calcific aortic valve stenosis as established from multiple epidemiological studies, including secondary analyses of clinical trials of patients treated with statins and proprotein convertase subtilisin/kexin 9 (PCSK9) inhibitors.^1–5^ Cardiovascular outcomes trials with selective Lp(a) lowering therapies are being conducted only in very high-risk ASCVD patients, but the high prevalence of elevated Lp(a) levels in patients with early onset ASCVD suggests the need for earlier intervention or primary prevention trials.

The contribution of risk-enhancing biomarkers (high-sensitivity C-reactive protein [hsCRP], coronary artery calcium [CAC]) and measures of subclinical atherosclerosis to enhance Lp(a) associated risk stratification in a primary prevention trial of selective Lp(a) lowering therapies is an important consideration in trial design.^6^

Coronary artery plaque characteristics are useful surrogates for ASCVD outcomes.^7^ Among patients undergoing coronary angiography for evaluation of coronary artery disease (CAD), elevated Lp(a) levels are associated with a more severe coronary phenotype that encompasses a higher prevalence of vulnerable plaques and increased coronary atheroma volume.^8–13^ Among patients undergoing coronary CT angiography (CCTA) for suspected CAD, several coronary plaque characteristics predicted a higher incidence of ASCVD events.^14^

Some analyses have shown that the increased risk of Lp(a)-associated cardiovascular events is dependent on a high systemic inflammatory state and potentially contributes to a high-risk coronary phenotype. Recent studies continue to show conflicting data with some in favor of hsCRP modification of Lp(a) and others showing no relationship between the two.^15–17^ According to recent guidelines, the use of CAC scoring is recommended for further risk assessment in cholesterol management, as it can independently predict ASCVD events.^18,19^ Similar to hsCRP, there has been conflicting evidence that CAC is associated with elevated Lp(a). The use of Lp(a) specific risk enhancers in a primary prevention cohort for clinical trials of selective Lp(a) lowering therapies is paramount to a successful trial. Non-calcified plaque volume has emerged as a potential target for coronary artery disease development.^20^

In this observational study, we investigated the associations between Lp(a) and coronary arterial plaque composition as measured by CCTA and the effect modification of CAC and hsCRP in a primary prevention cohort of 373 consecutive patients.

## METHODS

### Population

A total of 373 consecutive asymptomatic patients were evaluated for primary prevention of coronary artery disease at an outpatient cardiology clinic in New York, NY. All patients underwent CCTA imaging between 2018-2023. A retrospective analysis was performed on prospectively collected data. The study complied with the Declaration of Helsinki. The Institutional Review Board (IRB) of the Program for the Protection of Human Subjects at Icahn School of Medicine at Mount Sinai determined that the study (IRB ID: STUDY-24-00151) is not research involving human subjects as defined by the United States Department of Health and Human Services and Food and Drug Administration. The IRB of the Icahn School of Medicine at Mount Sinai waived ethical approval for this work.

### Laboratory measurements

Fasting blood samples were transported daily to the central laboratory (Cleveland HeartLab, Cleveland, OH) on wet ice for processing. Total cholesterol, high-density lipoprotein (HDL) cholesterol, and triglycerides were measured using the Beckman system and kits (Indianapolis, IN). Low-density lipoprotein (LDL) cholesterol was calculated using the Friedewald estimation. Lp(a) was measured in nmol/L with the Randox assay (Cleveland HeartLab, Cleveland, OH). High sensitivity hsCRP was measured using Abbott’s immunoturbidimetric assay (Cleveland HeartLab, Cleveland, OH). Lp(a) corrected LDL cholesterol was calculated by the Rosenson Marcovina equation.^21^

### CCTA acquisition

CCTA imaging was performed at two centers using a 128-detector row CT scanner (Somatom Definition AS+, Siemens Medical Solutions, Malvern, PA), and a 256-detector row scanner (Revolution Apex, GE HealthCare, Waukesha, WI). CCTA imaging was performed at two centers using a 128-detector row CT scanner (Somatom Definition AS+, Siemens Medical Solutions, Malvern, PA), and a 256-detector row scanner (Revolution Apex, GE HealthCare, Waukesha, WI). Sublingual nitroglycerin spray was administered to all patients without contraindication immediately prior to CCTA imaging. Patients were also given oral metoprolol an hour prior to examination, as needed, to achieve a target heart rate of <65 beats/min. Dosages of 50, or 100mg were chosen according to resting heart rate and BMI. CCTA parameters for the 128-slice scanner entailed a section collimation of 128•0.625mm, a gantry rotation time of 270ms, a tube current of 800 to 1,000 mA and 200 to 360mA (adjusting mA based on patient’s body size), and a tube voltage of 120 kV. CCTA parameters for the 256-slice scanner entailed a section collimation of 256•0.625mm, a gantry rotation time of 280ms, a tube current of 100-940mA, and a tube voltage of 100-120kV, both adjusted for patient’s body size. Prospective electrocardiogram–gated CCTA acquisition was applied, triggered at 75% of the R-R interval. For visualization of the coronary lumen, depending on location, a bolus of 80ml iohexol (Omnipaque 350, GE Healthcare, Waukesha, WI) or 65ml iohexol (Visapaque 320 GE Healthcare, Waukesha, WI) was injected intravenously (5.0 ml/s) followed by an immediate 50 ml saline bolus. Scans were triggered using an automatic bolus tracking technique, with a region of interest in the descending thoracic aorta.

### Atherosclerosis Imaging Quantitative CT

An AI-powered software solution (Cleerly Inc., Denver, CO) was used for the analysis of the CCTA images.^22^ This software utilizes a set of validated convolutional neural networks for tasks including evaluating image quality, segmenting, and labeling coronary structures, assessing lumen walls, determining vessel contours, and characterizing plaques. The AI-QCT system had been previously validated through comparisons with expert consensus, quantitative coronary angiography, fractional flow reserve, and intravascular ultrasound in multicenter trials, as previously documented in published studies.^22,23^

Initially, the algorithm generates centerline, lumen, and contour outlines for the outer vessel wall in each available phase. Subsequently, it identifies the most suitable series for detailed analysis. The decision regarding the highest-quality image is then made for each individual vessel. Once automated segmentation and labeling are completed for all vessels, plaques are assessed and measured using the HU attenuation. Lastly, a trained radiologic technologist with oversees the quality assurance of the AI analysis under the guidance of a CCTA qualified radiologist or cardiologist.

Coronary segments with a diameter ≥1.5 mm were incorporated into the analysis using the modified 18-segment model outlined by the Society of Cardiovascular Computed Tomography.^24^ The assessment of coronary percentage stenosis was determined for each vessel in accordance with the guidelines provided by the Society of Cardiovascular Computed Tomography.^25,26^ The categorization was done following the Coronary Artery Disease Reporting and Data System (CAD-RADS).^25^ Plaque was defined as any tissue structure >1 mm² within the coronary artery wall and distinguishable from the neighboring epicardial tissue, epicardial fat, or the vessel lumen itself.

Plaque volumes (mm³) were computed for each individual coronary lesion and then summed to determine the total plaque volume at the patient levels. Categorization of plaque volume involved the utilization of Hounsfield Unit (HU) ranges. Low-density noncalcified plaque (LD-NCP) was defined as plaques with any component on a pixel-level basis and quantified on an increment of 0.1μL as <30 HU. Noncalcified plaque volume (NCPV) was defined as HU values ranging from 30 to +350, while calcified plaque volume (CPV) comprised HU values >350 HU.^25^ To account for variations in coronary artery volume, coronary plaque volume was normalized relative to the total vessel volume per patient. This normalization was performed as follows: plaque volume/vessel volume×100%. The resultant normalized volumes were reported as percentage atheroma volume (PAV), percentage NCPV, and percentage CPV. Arterial remodeling was assessed by dividing the lesion diameter by the normal reference diameter. Positive remodeling was determined by a ratio ≥1.1.

Coronary lesions featuring both LD-NCP and positive remodeling were defined as two-feature-positive plaques.^26^ In cases where image quality was compromised due to factors such as motion, insufficient opacification, beam hardening, or other artifacts, only the section of the coronary artery affected by poor quality was excluded from the analysis.

### Statistical analysis

Data are presented as mean±standard deviation for normally distributed variables or median with interquartile range (IQR) for non-normally distributed variables. Categorical variables are expressed as number (percentage). To assess baseline and demographic differences in the high-risk plaque group versus non-high-risk plaque group, Student’s t-tests were used for normally distributed continuous data and Wilcoxon-Mann-Whitney test for non-normally distributed continuous data, while Chi-square or Fisher’s test were used for categorical variables. High-risk plaque was defined as the presence of low-density plaque (<30 Hounsfield units) with a Remodeling Index>1.1.^27^ All analyses were performed using SAS software version 9.4 (SAS Institute Inc., Cary, NC, USA). The data was split into quintiles using the SAS software that divided the population to have similar percentages in each group based on distribution (Supplemental Figure S1). While there were still significant associations when using Lp(a) as a continuous variable, using quintiles allowed for a more quantifiable increase in plaque volumes. A linear regression model was fit to assess the relationship between Lp(a) (≥125 nmol/L or <125 nmol/L), hsCRP (≥1mg/L or <1mg/L), and the different plaque volumes. In the multivariable analysis, the models were additionally adjusted for CAD risk factors, including age, sex, race, type 2 diabetes, hypertension, statin use, body mass index, smoking status, and calcium score. A two-sided p-value <0.05 was considered statistically significant.

## RESULTS

### Demographics

The mean age of the cohort was 56.2±8.9 years, 71.6% were male, 97.6% were white, and 54.2% were on statin therapy. Mean BMI was 26.4±4.5 kg/m^2^. (Table 1). The majority of patients were non-smokers (78.3%), roughly half of the patients had a family history of CAD as determined by a prior event (49.3%), hypertension (48.8%), non-calcified plaque 48.8%, and very few patients had type 2 diabetes (5.4%). Median LDL-C was 103(72,136)mg/dl, median Lp(a) was 31 (11, 89) nmol/L, median hsCRP levels were 0.8(0.4,1.8)mg/L, and median CAC score was 6.0(0.0,110.0). The Lp(a) corrected LDL-C median was 101(64, 131)mg/dL. Lp(a) quintiles were divided into 0-10, >10-20, >20-46, >46-112, >112. Table 1. Central illustration. Plaque volume distribution is presented in Table 2 and in Figure 1.

**Figure 1.**
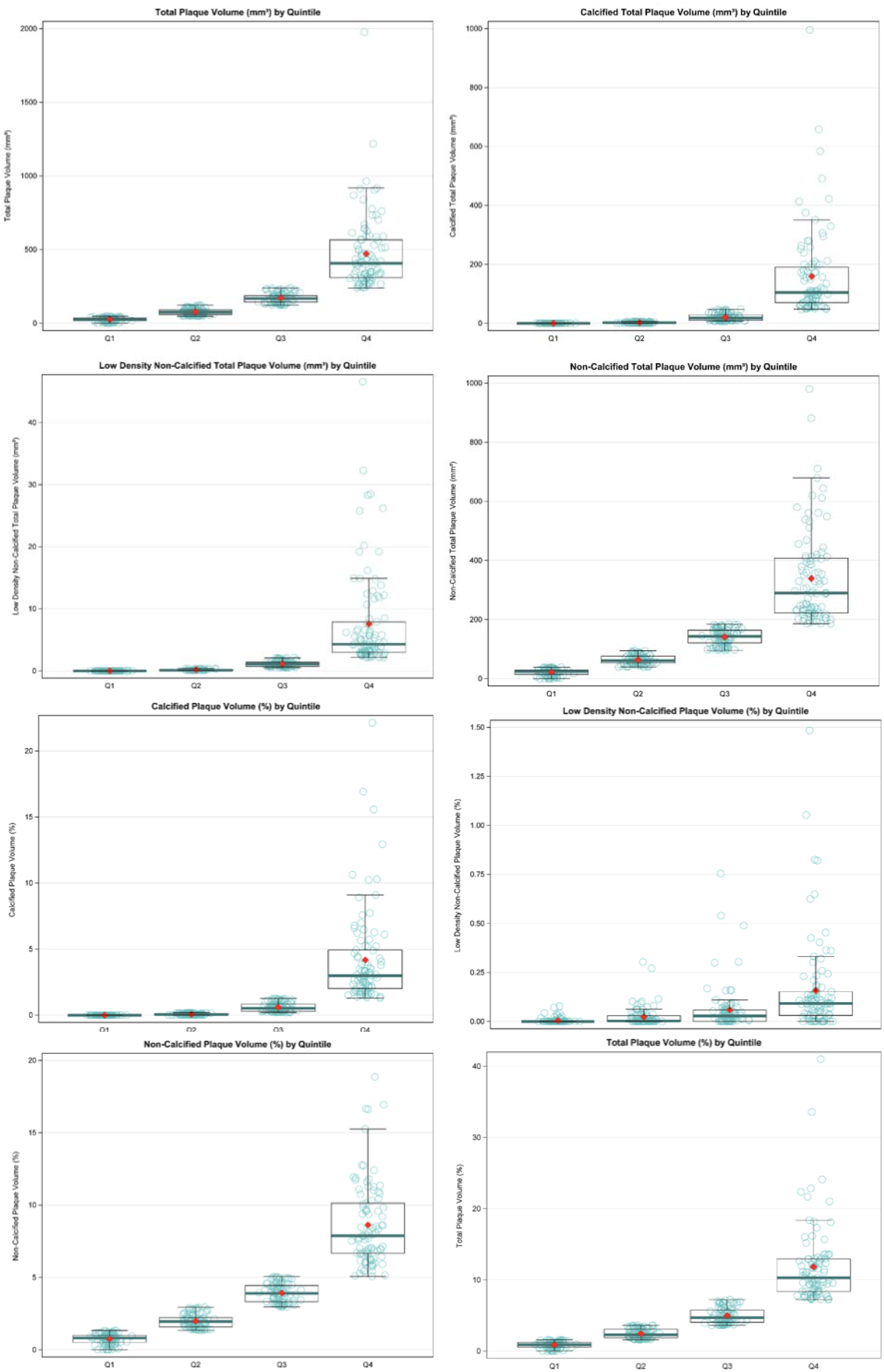
Box and Whisker Plots of Plaque Characteristics by Quintile

**Table 1:** Patient Demographics.

| **PATIENT DEMOGRAPHICS** |  |  |  |  |  |  |
| --- | --- | --- | --- | --- | --- | --- |
| Lp(a) Quintiles |  |  |  |  |  |  |
|  | Overall<br>population<br>N = 373 | Lp(a) 0-10<br>N =<br>90(24.0%) | Lp(a)<br>>10-20<br>N = 62<br>(17.0%) | Lp(a)<br>>20-46<br>N = 72<br>(19.0%) | Lp(a)<br>>46-112<br>N = 75<br>(20.0%) | Lp(a)<br>>112<br>N = 74<br>(20.0%) |
| Age, years | 56.2±8.9 | 55.1±9.2 | 56.2±8.7 | 56.6±9.2 | 55.4±9.1 | 58.1±8.2 |
| Sex, female | 106(28.4%) | 30(33.3%) | 14(22.6%) | 22(30.6%) | 17(22.7%) | 23(31.1%) |
| Race |  |  |  |  |  |  |
| Black | 1(0.3%) | 0(0.0%) | 0(0.0%) | 0(0.0%) | 0(0.0%) | 1(1.4%) |
| Other | 8(2.1%) | 4(4.4%) | 1(1.6%) | 2(2.8%) | 0(0.0%) | 1(1.4%) |
| White | 364(97.6%) | 86(95.6%) | 61(98.4%) | 70(97.2%) | 75(100.0%) | 72(97.3%) |
| BMI(kg/m<sup>2</sup>) | 26.4±4.5 | 27.2±5.4 | 26.7±4.7 | 26.2±4.2 | 26.3±4.2 | 25.7±3.8 |
| **LABORATORY DATA** |  |  |  |  |  |  |
| Calculated LDL cholesterol(mg/dL) | 103.0(72.0, 136.0) | 114.0(78.0, 139.0) | 115.5(70.0, 130.0) | 97.0(71.0, 124.0) | 105.0(71.0, 141.0) | 89.0(66.0, 134.0) |
| Apolipoprotein B(mg/dL) | 85.0(66.0, 105.0) | 89.0(70.0, 107.0) | 84.0(63.0, 97.0) | 79.0(67.0, 91.0) | 86.5(68.0, 109.0) | 82.0(65.0, 113.0) |
| HDL cholesterol(mg/dL) | 59.0(50.0, 71.0) | 58.5(49.0, 69.0) | 58.5(51.0, 73.0) | 59.5(47.0, 75.0) | 58.0(52.0, 72.0) | 60.5(52.0, 70.0) |
| Total Cholesterol(mg/dL) | 183.0(148.0, 218.0) | 191.0(159.0, 223.0) | 189.5(147.0, 214.0) | 176.0(153.5, 210.5) | 183.0(150.0, 223.0) | 174.0(140.0, 230.0) |
| Triglycerides(mg/dL) | 81.0(64.0, 113.0) | 90.0(68.0, 117.0) | 79.5(62.0, 113.0) | 75.0(60.5, 104.5) | 78.0(62.0, 115.0) | 76.5(66.0, 110.0) |
| Lp(a)(nmol/L) | 31.0(11.0, 89.0) | 10.0(10.0, 10.0) | 15.0(12.0, 18.0) | 32.0(25.0, 38.0) | 74.0(54.0, 89.0) | 185.5(152.0, 265.0) |
| Lp(a)-corrected LDL cholesterol(mg/dL) | 101.2(64.1, 131.2) | 113.2(77.2, 138.2) | 114.3(68.9, 129.2) | 94.9(68.6, 121.3) | 99.2(65.4, 135.3) | 67.9(51.1, 118.2) |
| High-sensitivity C-reactive protein(mg/L) | 0.8(0.4, 1.8) | 1.0(0.5, 2.1) | 0.7(0.4, 1.4) | 1.1(0.4, 1.9) | 0.7(0.4, 1.8) | 0.9(0.4, 1.5) |
| **CLINICAL CHARACTERISTICS** |  |  |  |  |  |  |
| CAC Score | 6.0(0.0, 110.0) | 1.0(0.0, 56.5) | 34.0(0.0, 180.0) | 4.0(0.0, 116.0) | 4.0(0.0, 93.0) | 20.0(0.0, 137.0) |
| Diabetes Type 2 | 20(5.4%) | 3(3.3%) | 8(12.9%) | 3(4.2%) | 1(1.3%) | 5(6.8%) |
| Family History of CAD | 184(49.3%) | 38(42.2%) | 34(54.8%) | 37(51.4%) | 33(44.0%) | 42(56.8%) |
| Smoker |  |  |  |  |  |  |
| Former | 68(18.2%) | 18(20.0%) | 11(17.7%) | 15(20.8%) | 10(13.3%) | 14(18.9%) |
| No | 292(78.3%) | 68(75.6%) | 50(80.6%) | 56(77.8%) | 60(80.0%) | 58(78.4%) |
| Yes | 13(3.5%) | 4(4.4%) | 1(1.6%) | 1(1.4%) | 5(6.7%) | 2(2.7%) |
| Statin Use | 202(54.2%) | 42(46.7%) | 28(45.2%) | 36(50.0%) | 44(58.7%) | 52(70.3%) |
| Anti-inflammatory use (Chronic NSAID) | 11(2.9%) | 3(3.3%) | 2(3.2%) | 0(0.0%) | 3(4.0%) | 3(4.1%) |
| PCSK9i PCSK9 mAb Use | 9(2.4%) | 2(2.2%) | 1(1.6%) | 1(1.4%) | 4(5.3%) | 1(1.4%) |
| Anti-hypertensive use | 121(32.4%) | 28(31.1%) | 21(33.9%) | 23(31.9%) | 26(34.7%) | 23(31.1%) |
| Aspirin use | 62(16.6%) | 12(13.3%) | 14(22.6%) | 14(19.4%) | 8(10.7%) | 14(18.9%) |

**Table 2:** Plaque Volume Distribution.

| **PLAQUE VOLUME DISTRUBUTION** |  |
| --- | --- |
| Presence of High-Risk Plaque(Yes) | 182(48.8%) |
| # High Risk Plaques; Median(Q1, Q3) | 0.0(0.0,2.0) |
| Lesion Length; Median(Q1, Q3) | 61.0(29.5,136.3) |
| TPV(mm<sup>3</sup>); Median(Q1, Q3) | 122.6(47.9,239.7) |
| TCPV(mm<sup>3</sup>); Median(Q1, Q3) | 6.6(0.1,47.3) |
| TNCPV(mm<sup>3</sup>); Median(Q1, Q3) | 94.6(39.0,184.1) |
| LD-NCPV; Median(Q1, Q3) | 0.4(0.0,2.1) |
| PAV(%); Median(Q1, Q3) | 3.6(1.6,7.2) |
| PNCAV(%); Median(Q1, Q3) | 3.0(1.4,5.1) |
| PCAV(%); Median(Q1, Q3) | 0.2(0.0,1.3) |
| PLD-NCAV(%); Median(Q1, Q3) | 0.0(0.0,0.1) |
| Total Lumen Volume; Mean±Std | 3293.3±970.5 |
| Total Vessel Length(mm); Mean±Std | 626.0±100.8 |
| Total Vessel Volume(mm<sup>3</sup>); Mean±Std | 3480.2±1045.3 |
| # of Stenoses; Median(Q1, Q3) | 7.0(5.0,10.0) |
| Maximum Stenosis Diameter(%); Median(Q1, Q3) | 0.2(0.1,0.3) |
| Maximum Stenosis Area(%); Median(Q1, Q3) | 0.3(0.2,0.5) |
| Remodeling Index; Median(Q1, Q3) | 1.3(1.2,1.4) |
| Minimum Lumen Diameter of Smallest Stenosis(mm); Median(Q1, Q3) | 2.1(1.8,2.4) |

The high-risk plaque group had a statistically significantly higher BMI, and more patients were taking aspirin and statins (all p<0.05).

### Association between Lp(a) and coronary plaque characteristics

After adjustment for CAD risk factors, including age, sex, race, type 2 diabetes, hypertension, smoking status, statin use, and body mass index, every increase in Lp(a) quintile was associated with a 0.4% increase in LD-NCPV (P=0.039) and a 0.01% increase in PLD-NCAV (P=0.013). Lp(a) remained significantly associated with PLD-NCAV when analyzed as a continuous variable (P = 0.025). Increases in Lp(a) by quintiles and continuously were not associated with increases in TPV, TCPV, TNCPV, PAV, PCAV, PNCAV (P > 0.05). Table 3. Figure 2.

**Figure 2.**
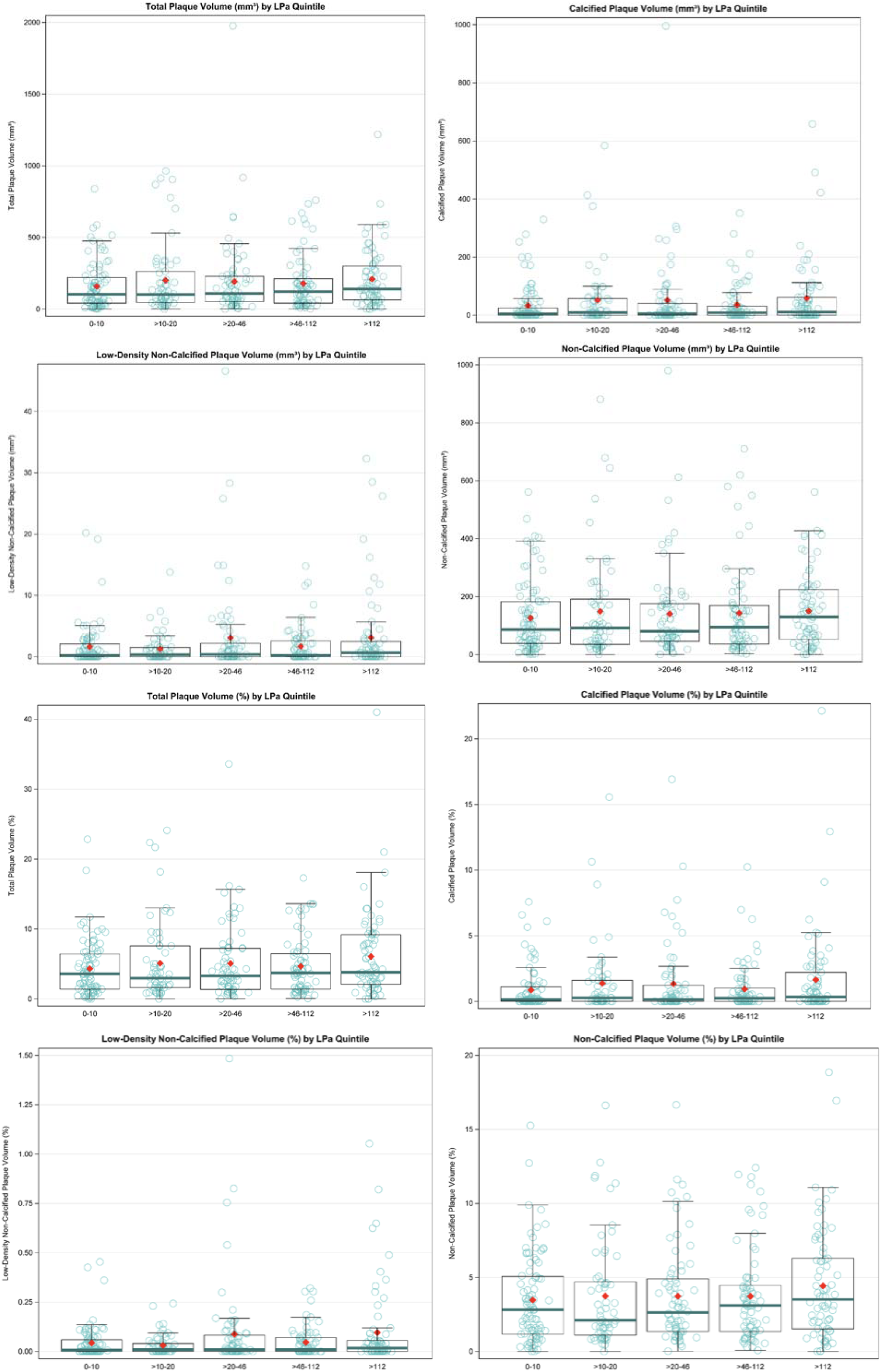
Box and Whisker Plots of Plaque Characteristics by Lp(a) Quintile

**Table 3:** Univariate and multivariate analysis for Lp(a) by quintiles.

| Outcome | Unadjusted |  | Adjusted* |  |
| --- | --- | --- | --- | --- |
|  | Beta Estimate | P-value | Beta Estimate | P-value |
| TPV(mm<sup>3</sup>) | 8.548 | 0.2631 | -0.536 | 0.9404 |
| TCPV(mm<sup>3</sup>) | 3.902 | 0.2669 | -1.418 | 0.6760 |
| TNCPV(mm<sup>3</sup>) | 4.646 | 0.3700 | 0.882 | 0.8565 |
| LD-NCPV | 0.330 | 0.0627 | 0.375 | 0.0387 |
| PAV(%) | 0.312 | 0.0805 | 0.081 | 0.6323 |
| PCAV(%) | 0.123 | 0.1612 | -0.020 | 0.8168 |
| PNCAV(%) | 0.189 | 0.1127 | 0.101 | 0.3791 |
| PLD-NCAV(%) | 0.012 | 0.0250 | 0.013 | 0.0137 |
| *Multivariate Model is not adjusted for CAC |  |  |  |  |

### Association between Lp(a) and CAC

There was no association between Lp(a) and CAC as analyzed continuously or by quintiles (P > 0.05).

### The effect of CAC on the association between Lp(a) and plaque characteristics

After adjustment for CAD risk factors and CAC, every increase in Lp(a) quintile was associated with a 0.2% increase in PAV (P=0.049), a 0.4% increase in LD-NCPV (P=0.046), and a 0.01% increase in PLD-NCAV (P=0.017). Table 4. However, there was no association between Lp(a) and CAC (P=0.631).

**Table 4:** Univariate and multivariate analysis for Lp(a) by quintiles, adjusting for CAC.

| Dependent Variable | Unadjusted |  | Adjusted* |  |
| --- | --- | --- | --- | --- |
|  | Beta Estimate | P-value | Beta Estimate | P-value |
| TPV(mm<sup>3</sup>) | 6.154 | 0.1686 | 4.717 | 0.2739 |
| TCPV(mm<sup>3</sup>) | 2.547 | 0.0233 | 1.532 | 0.1812 |
| TNCPV(mm<sup>3</sup>) | 3.607 | 0.4062 | 3.185 | 0.4475 |
| LD-NCPV | 0.326 | 0.0688 | 0.368 | 0.0458 |
| PAV(%) | 0.262 | 0.0140 | 0.209 | 0.0486 |
| PCAV(%) | 0.093 | 0.0049 | 0.055 | 0.0986 |
| PNCAV(%) | 0.169 | 0.0986 | 0.154 | 0.1300 |
| PLD-NCAV(%) | 0.012 | 0.0265 | 0.013 | 0.0172 |

## Association between CAC and coronary plaque characteristics

CAC is significantly associated with an increase in multiple plaque characteristics, including TPV, TCPV, TNCPV, PAV, PNCAV, PCAV (P<0.001). After adjustment for CAD risk factors and Lp(a), all associations remain statistically significant (P < 0.001) with similar beta estimates. Thre is no association between CAC and LD-NCPV and LD-NCPAV (P > 0.05). Table 5.

**Table 5:** Univariate and multivariate analysis for CAC.

| Dependent Variable | Unadjusted |  | Adjusted* |  |
| --- | --- | --- | --- | --- |
|  | Beta Estimate | P-value | Beta Estimate | P-value |
| TPV(mm<sup>3</sup>) | 0.589 | <.0001 | 0.557 | <.0001 |
| TCPV(mm<sup>3</sup>) | 0.316 | <.0001 | 0.308 | <.0001 |
| TNCPV(mm<sup>3</sup>) | 0.274 | <.0001 | 0.249 | <.0001 |
| LD-NCPV | 0.000 | 0.9502 | -0.000 | 0.9579 |
| PAV(%) | 0.014 | <.0001 | 0.013 | <.0001 |
| PCAV(%) | 0.008 | <.0001 | 0.008 | <.0001 |
| PNCAV(%) | 0.006 | <.0001 | 0.005 | <.0001 |
| PLD-NCAV(%) | -0.000 | 0.4497 | -0.000 | 0.5486 |

### Association between hsCRP and coronary plaque characteristics

After adjustment for CAD risk factors and CAC, for every quintile increase in hsCRP, there was no significant increase in PAV, TCPV, LD-NCPAV, or PCAV (P > 0.05). Table 6.

**Table 6:** Univariate and multivariate analysis for hsCRP by quintiles.

| Dependent Variable | Unadjusted |  | Adjusted* |  |
| --- | --- | --- | --- | --- |
|  | Beta Estimate | P-value | Beta Estimate | P-value |
| TPV(mm<sup>3</sup>) | 8.710 | 0.0622 | 5.127 | 0.2866 |
| TCPV(mm<sup>3</sup>) | -1.150 | 0.3288 | 0.580 | 0.6504 |
| TNCPV(mm<sup>3</sup>) | 9.860 | 0.0295 | 4.546 | 0.3311 |
| LD-NCPV | 0.170 | 0.3644 | -0.033 | 0.8746 |
| PAV(%) | 0.181 | 0.1052 | 0.144 | 0.2222 |
| PCAV(%) | -0.043 | 0.2136 | 0.013 | 0.7205 |
| PNCAV(%) | 0.224 | 0.0364 | 0.131 | 0.2476 |
| PLD-NCAV(%) | 0.006 | 0.2753 | 0.001 | 0.8264 |
| *Multivariate Model is not adjusted for CAC |  |  |  |  |

### The effect of hsCRP on the association between Lp(a) and plaque characteristics

There were no significant interactions between Lp(a) and hsCRP for TPV, TCPV, TNCPV, LD-NCP, PAV, PCAV, PNCAV, PLD-NCAV (P > 0.05).

## DISCUSSION

In this single-center study of 373 primary prevention patients undergoing clinically indicated CCTA for improved risk stratification, elevated Lp(a) was associated with an increase in low-density non-calcified plaque volume. Non-calcified plaque is an imaging plaque characteristic associated with the highest risk of subsequent cardiovascular events.^14,28,29^ We found no significant association between CAC and Lp(a); however, CAC modified the association between Lp(a) and percent atheroma volume. Additionally, there was no association between hsCRP and plaque nor did hsCRP impact the association between Lp(a) and coronary plaque. These data confirm the relation of Lp(a) with coronary atherosclerosis, but do not demonstrate the utility of CAC or hsCRP on high-risk coronary plaque features.

This is the first study to analyze plaque volumes and characteristics in an asymptomatic primary prevention cohort. Compared to symptomatic patients with high suspicion for CAD, this cohort has lower TPV, LD-NCPV, PAV, and PLD-NCAV.^23,30^ Additionally, this cohort has a higher amount of non-calcified than calcified plaque volumes (median NCPV 94.6 vs. CPV 6.6), This is likely attributable to the younger age of the population, as their plaques have not yet had sufficient time to calcify.

Previous studies have investigated the relationship between Lp(a) and coronary plaque characteristics in patients with advanced coronary artery disease. A post-hoc analysis of six randomized trials using intravascular ultrasound (IVUS) to characterize coronary atheroma found that elevated Lp(a) is independently associated with an increased percent atheroma volume.^8^ Additionally, two studies using optical coherence tomography (OCT)^9,10^ found that in patients with acute coronary syndrome (ACS), those with higher Lp(a) levels had increased atherosclerotic burden and an increased prevalence of thin cap fibroatheroma. Near-infrared spectroscopy (NIRS) has also been used to demonstrate that in patients with CAD, those with lower Lp(a) had coronary lesions with a smaller amount of lipidic plaque.^31^ IVUS, OCT, and NIRS are all invasive modalities used for plaque quantification and characterization, of which IVUS is the current gold standard.^32^ A recent study using CCTA, a non-invasive alternative, showed that in patients with advanced CAD, those with higher Lp(a) had an accelerated progression of low-density plaque.^11^

Our study, using CCTA in a population without known CAD, demonstrated that Lp(a) is associated with low-density plaque. In primary and secondary prevention contexts, it has been shown that elevated levels of Lp(a) can predict the incidence of ASCVD^12^ and increase the risk of MI, especially in patients with low-density non-calcified plaque.^33^ It is possible that the increase in low-density non-calcified plaque, a high-risk feature plaque phenotype, may increase cardiovascular risk.^11^ This study further supports using plaque characterization as an anatomical predictor of events and the need for further analysis between Lp(a) and plaque characterization.

Multiple studies, including the SCOT-HEART (Scottish Computed Tomography of the Heart), ICONIC (Incident Coronary Syndromes Identified by Computed Tomography), and PROMISE (Prospective Multicenter Imaging Study for Evaluation of Chest Pain), have shown that both quantitative and qualitative evaluations of atherosclerosis offer predictive value for future ASCVD events.^34–36^ These coronary imaging characteristics included plaque location, extent, and high-risk plaque characteristics. Yang et al. reported similar findings that included six features of the lesions that were most clinically relevant in providing improved prognostication, which included percent atheroma volume, plaque volume (in this study, total plaque volume), fibrofatty necrotic core (in this study, low-density non-calcified plaque), minimal lumen area, remodeling index, and location in the proximal LAD.^14^

The implementation of artificial intelligence–guided quantitative coronary computed tomography angiography analysis (AI-QCT) has facilitated an expedited assessment of atherosclerotic plaque characteristics and burden.^22,37^ Many studies demonstrate that AI-QCT analysis is objective, reproducible, and able to achieve diagnostic accuracy in detecting obstructive stenosis and quantifying atherosclerotic plaque burden.^23^

In this asymptomatic cohort, we assessed risk enhancers, such as hsCRP and CAC, to better evaluate the risk of ASCVD related to Lp(a). Studies have shown that hsCRP modifies the association between Lp(a), and cardiovascular events. ACCELERATE (Assessment of Clinical Effects of Cholesteryl Ester Transfer Protein Inhibition With Evacetrapib in Patients at a High-Risk for Vascular Outcomes), MESA (Multi-Ethnic Study of Atherosclerosis), and REGARDS (Reasons for Geographic and Racial Differences in Stroke) demonstrated that increased levels of Lp(a) are associated with cardiovascular death, myocardial infarction, and stroke only when hsCRP levels are equal to or greater than 2mg/L,^38–40^ while others showed a link only when hsCRP levels were above 4.2mg/L.^41^ Others have shown that elevated levels of Lp(a) are a significant independent factor contributing to the risk of ASCVD irrespective of hsCRP levels.^16,42^ In a recent Biomar-CaRE (Biomarker for Cardiovascular Risk assessment across Europe) project, hsCRP only modified risk in participants with known coronary heart disease (CHD) when hsCRP≥2mg/L.^15^ In those without CHD, increased Lp(a) levels were associated with incident CHD irrespective of hsCRP concentration. It has also been shown that only after adjusting for the usual cardiovascular risk factors, hsCRP is just weakly associated with the presence of coronary atherosclerosis.^43^ The findings in the current study, a primary prevention cohort, support the latter, that even in the absence of elevated hsCRP levels, Lp(a) levels are associated with increased plaque volume and a high-risk plaque phenotype. This is possibly due to the overall low levels of hsCRP among the current population, however, 20.6% of patients had hsCRP levels ≥2mg/L. Therefore, elevated levels of hsCRP may be a risk enhancer for cardiovascular events but are not associated with coronary artery plaque characteristics. These findings suggest that among patients for primary prevention, a more in-depth characterization of Lp(a) associated cardiovascular risk is necessary, compared to the conventional risk enhancers such as hsCRP and CAC, which may include using CCTA as a screening tool as Lp(a) is often accompanied by severe adverse cardiovascular outcomes in young adults.

According to recent guidelines, the American College of Cardiology and American Heart Association recommend using CAC scoring for further risk assessment in cholesterol management, as it can independently predict ASCVD events.^18,19^ Similar to hsCRP, there has been conflicting evidence that CAC is associated with elevated Lp(a). It was originally shown that there was no association between Lp(a) and CAC in the Dallas Heart Study (DHS).^44^ More recently an analysis of MESA and DHS showed that Lp(a) and CAC were independently associated with ASCVD and when both were elevated, patients were at a significantly higher risk of ASCVD.^45^ In this patient population, median CAC was 6, which places most of these patients between the 25^th^ percentile (CAC=0) and 50^th^ (CAC=9) for age, sex, and gender.^46^ Our study found no significant association between Lp(a) and CAC. Interestingly, the current study showed that CAC was independently associated with total plaque, calcified and non-calcified plaque. However, there was no association between CAC and low-density non-calcified plaque, whereas Lp(a) was independently associated with high-risk low-density non-calcified plaque. Therefore, the present study demonstrates the importance of using plaque composition as risk stratification compared to CAC.

Several professional society scientific statements recommend testing Lp(a) at least once in adults.^1,2,47,48^ Despite current existing therapies with statins and PCSK9i, elevated Lp(a) continues to increase risk of ASCVD, mainly due to inflammation and oxidized phospholipids.^6^ While clinical outcomes trials of Lp(a) lowering therapies with antisense oligonucleotides (HORIZON NCT04023552) and small interfering RNA (OCEAN(a) NCT04270760) trials are underway, evidence supporting Lp(a) as a target of therapy awaits completion of the phase 3 trials.^49,50^ Future trial design still remains a challenge in order to optimize outcomes and patient selections.^6^ While the current trials focus on high-risk populations, our data helps support the need for a primary prevention trial given that Lp(a) is an inherited risk factor and elevated levels lead to early-onset CAD and currently the conventional measure of CAC is not adequate for risk stratification. It also further supports the use of plaque characteristics to better assess the effects of Lp(a) lowering therapies.

Limitations of this study include the fact that the patient population was from a single center that is skewed in the relative uniformity of its participants. Additionally, the differences between the two scanners may have affected plaque thresholds. It is important for future research to investigate the association between Lp(a), inflammatory markers, and plaque characteristics in a larger, more diverse population, and higher distributions of Lp(a) concentrations. There may be many implications of this research. Large randomized controlled trials using Lp(a) targeting drugs could use AI-QCT to assess baseline plaque characteristics as inclusion criteria for a primary prevention population, and further serve as an outcome in order to investigate the efficacy of Lp(a) lowering on coronary plaque characteristics that have been shown in longitudinal studies to predict major adverse cardiovascular outcomes.

## Data Availability

All data produced in the present study are available upon reasonable request to the authors.

## Acknowledgments

n/a

## Supplemental Material

Figure S1.

## Non-standard Abbreviations and Acronyms

Lp(a): Lipoprotein(a)
CAD: coronary artery disease
ASCVD: atherosclerotic cardiovascular disease
hsCRP: high sensitivity C-reactive protein
CAC: coronary artery calcium
AI-QCT: artificial intelligence quantitative coronary CTA
CCTA: coronary CT angiography

---

**Central Illustration.**
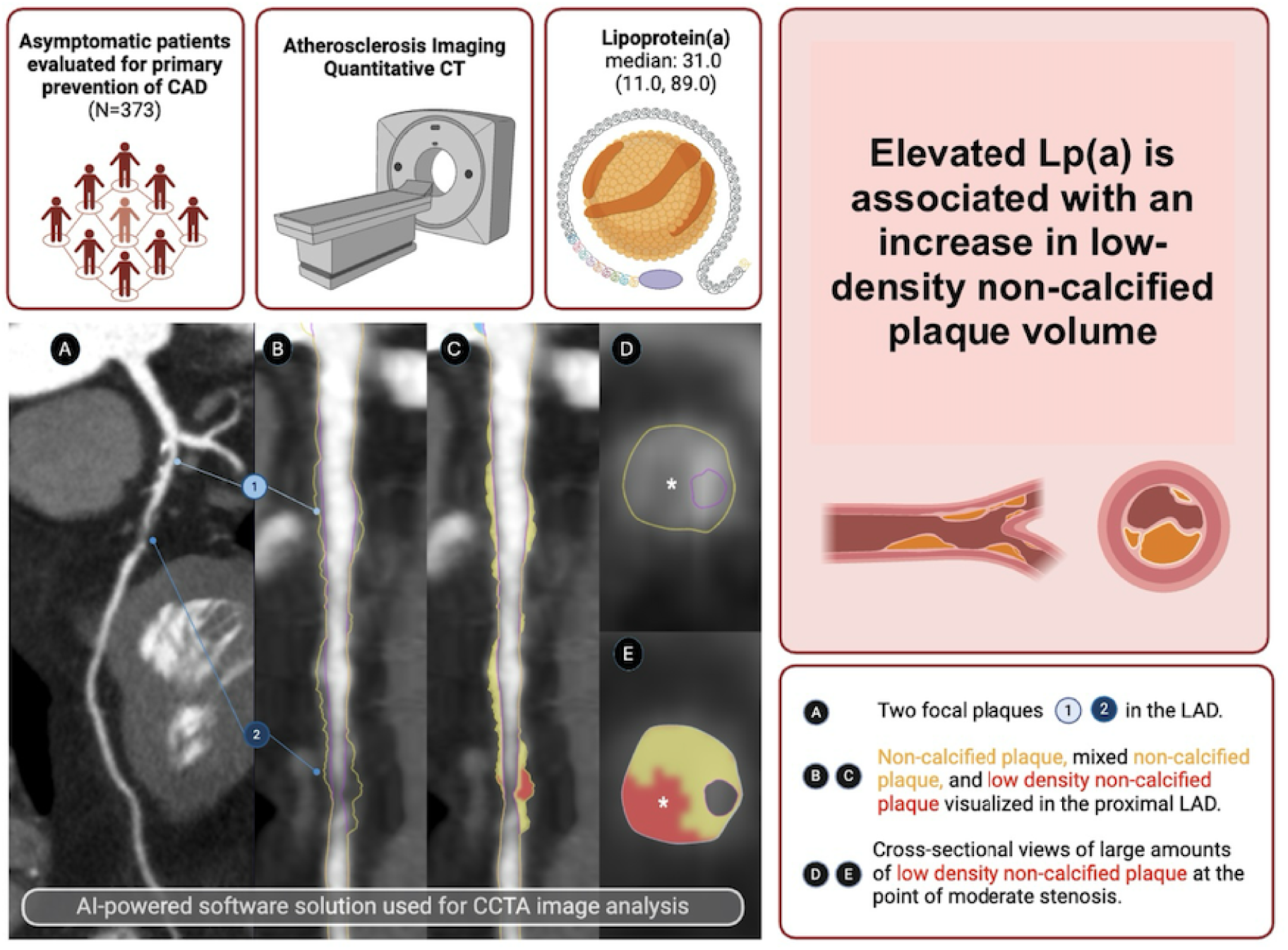
Evaluation of the relationship between lipoprotein(a) and coronary plaque characteristics 373 consecutive asymptomatic patients were evaluated for primary prevention of CAD. AI-QCT was used to investigate the relationship between Lp(a) and coronary plaque characteristics. In this cohort, elevated Lp(a) concentrations was significantly associated with increased total coronary plaque burden and particularly, low-density non-calcified plaque volume.

