## Supplemental Figure S1 for "Lipoprotein(a) is Associated with Increased Low-Density Plaque Volume"

**Figure S1.**
Distribution and n's for each quintile.

| Rank for Variable by Lp(a) | | |
|---|---|---|
| Lp(a) rank | Frequency | Percent |
| 0-10 | 90 | 24.13 |
| >10-20 | 62 | 16.62 |
| >20-46 | 72 | 19.30 |
| >46-112 | 75 | 20.11 |
| >112 | 74 | 19.84 |

Histogram of Lp(a)

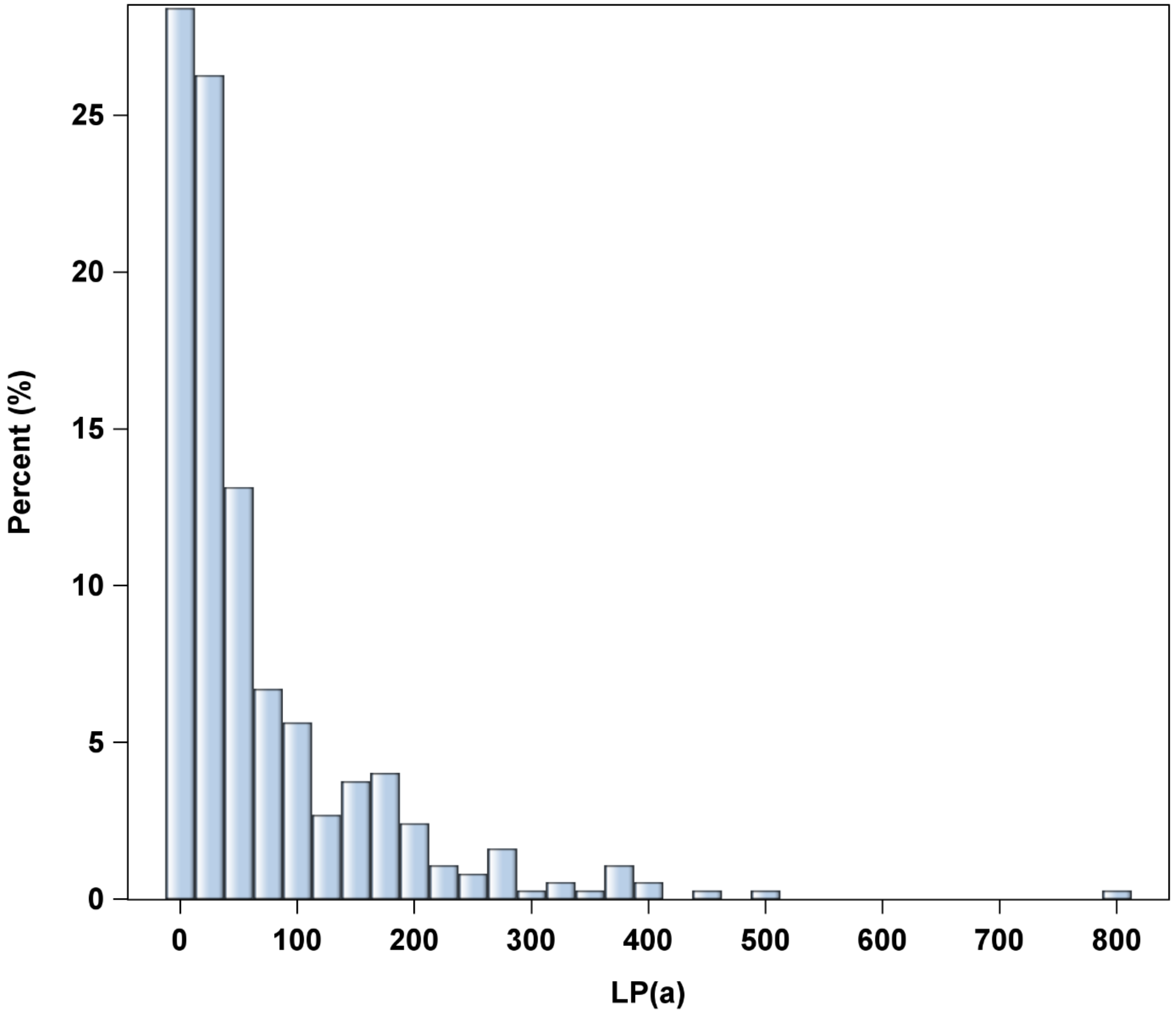
